# Effects of metformin use in pregnant women on maternal and infant outcomes: protocol for a systematic review and meta-analysis

**DOI:** 10.1101/2022.08.13.22278738

**Authors:** Kezhang Wang, Rui Pan, Yu Wang

## Abstract

**Introduction:** Metformin has been considered for use in women who are overweight or obese and with pre-existing type 2 diabetes(T2DM) or gestational diabetes mellitus(GDM). Nevertheless, few studies have weighed the pros and cons of maternal metformin use on offspring and women’s perinatal health. We determined to ascertain whether the insulin sensitiser metformin improves fetal and maternal outcomes in Pregnant women with type 2 diabetes or with a BMI no less than 24 kg/m^2^

**Methods and analysis:** The protocol was formulated according to Preferred Reporting Items for Systematic Review and Meta-Analysis Protocols (PRISMA-P). Exhaustive literature searches of electronic bibliographic databases will be conducted in PubMed, EMBASE, Cochrane Library, Web of Sciences, Scopus, ClinicalTrial, OvidSP and ScienceDirect. We used a predefined data extraction table to perform data extraction, and publication bias measurement for each study will be accomplished through the Cochrane Collaboration Tool.

The statistical analyses will be performed using RevMan (5.4.1; The Cochrane Collaboration) and STATA (17). Heterogeneity will be assessed using the Q and I^2^ test, with a value of I^2^ > 50% being considered substantial. The extent of publication bias will be investigated by visual assessment of a funnel plot and using Egger’s test, Begg’s test, and Harbord’s test if ten or more trials are pooled.

**Ethics and dissemination:** Patients’ consent and Ethical approval are not requisites. The complete analysis will be published in an international open-access journal and translational articles to disseminate the results to clinical internists.

PROSPERO registration number: CRD42022347084

**Strengths and limitations of this study:**

▸ This review will comprehensively assess published English-language studies without period or geographical restrictions.
▸ This system review plan strictly follows preferred reporting items for systematic review and meta-analysis protocols (PRISMA-P)
▸ Included studies will be limited to RCTs, which have a high degree of internal consistency and are not representative of the real world.
▸ As is the case with most meta-analyses, there may be significant and unexplained heterogeneity in this study.

## INTRODUCTION

The morbidity of GDM and pregnant women with pre-existing T2DM has notably increased in recent decades. One etiology can be attributed to an increased maternal overweight or obesity incidence.^1-2^ The steady climb in the global incidence of GDM may also result from declining criteria for diagnosis, new screening methods, or other increases in population risk factors aside from obesity.^3-4^ Women with GDM during pregnancy or pre-existing T2DM have increased occurrence rates of adverse outcomes, including malformations, cesarean section, preeclampsia, macrosomia, preterm birth, birth injury, shoulder dystocia, and large for gestational age (LGA).^5-6^ Medical nutrition therapies (MNT), used in clinical practice, including food planning, dietary shifts, lifestyle rectification, and more physical activity, are regarded as the foundation of treatment for GDM.^7^ Nevertheless, therapy with extra insulin is conventionally deemed the first-line medical treatment for mothers with refractory hyperglycemia with MNT.^8-10^In Europe, most women with pre-existing T2DM are treated with insulin analogs before and during pregnancy. Additionally, some of them are using insulin pumps during pregnancy, and the application of constant glucose monitoring is becoming more conventional.^6^

Being overweight or obese jeopardises no less than 1.9 billion adults, 41 million children below the age of 5 years, and 270 million children aged 5 to 17 years,^11^ worldwide. Overweight is an individual with a body mass index (BMI) between 25.0 kg/m2 and 29.9 kg/m^2^. In comparison, obesity is defined as a BMI of 30.0 kg/m2 or more (In China, 24.0 kg/m^2^ and 28.0 kg/m^2^, respectively). Over 50% of pregnant women in the US are defined as overweight or obese. Infants born to these women are at elevated risk of hypoglycemia, prematurity, stillbirth, admission to the neonatal intensive care unit, respiratory distress at birth, childhood obesity, macrosomia with a possible birth injury, and congenital anomalies. Overweight or obese women are also at an elevated risk of complications in pregnancy.^12^ It can be concluded that maternal overweight or obesity is also closely related to adverse outcomes on the long-term health of infants, including the risk of cardiovascular disease, overweight, and diabetes. In light of the massive, hazardous implications of maternal overweight or obesity, international guidelines suggest no less than 30 minutes of moderate-intensive physical activity each day, including aerobic and resistance training.^13^

There is substantial evidence indicating that exercise could have a preventive effect on the initiation and development of fetal cesarean sections and macrosomia.^14-15^ Metformin (N, N-dimethyl biguanide), a biguanide oral antidiabetic medication, has received extensive recognition. Metformin is authorised for application in treating GDM in many countries worldwide. It is enrolled in the 20th World Health Organization (WHO) essential medicines list. The Society has classified it for Maternal-Fetal Medicine (SMFM) as one of the GDM’s first-line treatments.^16^ Considerable evidence indicates that metformin efficiently stabilises maternal glycaemic control and might be conducive to curbing gestational weight gain.^17^ Nevertheless, there are latent safety concerns that metformin can go through the placenta, which might directly influence fetal physiology homeosis.^18-19^ Other studies of women with GDM have found elevated risks of small for gestational age (SGA) and premature delivery.^20-21^ Simultaneously, more and more pregnant women are being exposed to metformin, whereas knowledge of the medication’s short-term and long-term effects remains insufficient.^22^ Although previous systematic reviews or meta-analyses have compared metformin to insulin or metformin alone used in diabetic or obese pregnant women, the quality and quantity of randomised control trials(RCT) were limited. Several high-quality, high sample content RCTs have been published since then. Therefore, our study aimed to evaluate the pros and cons of metformin as a treatment for pregnant women with pre-existing T2DM or GDM and pregnant women whose pre-pregnancy BMI is above 25.0 or 30.0. Unlike the previous studies, which focus on metformin’s effect on women’s perinatal health, we mainly concentrate on metformin’s implications for offspring’s health. Resolving this issue is significant as the number of pregnant women receiving metformin increases worldwide.

## METHODS AND ANALYSIS

### Protocol and registration

We conceived this protocol following preferred reporting items for systematic review and meta-analysis protocols (PRISMA-P). By the guidelines, this study protocol has been registered on the International Prospective Register of Systematic Reviews (PROSPERO registration number: CRD42022347084)

### Search method for identification of studies

Exhaustive literature searches of electronic bibliographic databases will be conducted in PubMed, EMBASE, Cochrane Library, Web of Sciences, Scopus, ClinicalTrial, OvidSP, and ScienceDirect. For the rarity of existing literature, we placed no time restrictions on the date of publication.

Exhaustive literature searches of electronic bibliographic databases will be conducted from inception to the present day in PubMed, EMBASE, Cochrane Library, Web of Science, Scopus, ClinicalTrials, OvidSP, and ScienceDirect. We did not restrict the types, dates, and statuses of the publications eligible for inclusion. The search strategy was composed of the following group terms:

#1 “metformin” [Mesh]

#2 metformine*[Title/Abstract] OR “LA-6023” [Title/Abstract] OR LA6023[Title/Abstract] OR Dimethylbiguanidine*[T itle/Abstract] OR Dimethylguanylguanidi ne*[Title/Abstract] OR Glucophage*[Title/Abstract] OR Metformiinihydrokloridi *[Title/Abstract] OR Metforminhidroklorid*[Title/Abstract] OR Metforminhydroklorid* [Title/Abstract]

#3 (Metformini*[Title/Abst ract] OR Metformino*[Title/Abst ract]) AND (hidrochlorida*[Title/A bstract] OR hydrochloridum*[Title/ Abstract])

#4 #1 OR #2 OR #3

#5 “pregnancy” [Mesh]

#6 pregnancies*[Title/Abstract] OR Gestation*[Title/Abstract]

#7 (prenatal*[Title/Abstract] OR antenatal*[Title/Abstract]) AND (care*[Title/Abstract])

#8 “pregnant women” [Title/Abstract] OR “pregnant woman” [Title/Abstract]

#9 #5 OR #6 OR #7 OR #8

#10 randomized controlled trial[Publication Type] OR random*[Title/Abstrac t] OR placebo[Title/Abstract]

#11 #4 AND #9 AND #10

#12 #4 AND #9 AND #10 NOT (“polycystic ovarian syndrome”[Title/Abstract] OR”PCOS” [Title/Abstract])

### Criteria for considering studies to review

Studies will be included if they satisfy these eligibility criteria:

① **Type of participants**: eligible participants are pregnant women (between 6 and 22 weeks plus six days’ gestation) with type 2 diabetes (fasting blood glucose ≥ 7.0mmol/L, or random blood glucose ≥11.1mmol/L, or two h after OGTT blood glucose ≥ 11.1mmol/L) or with a BMI of 24 kg/m^2^ or more. And the BMI is pre-pregnancy BMI. Trials including pregnant women with polycystic ovary syndrome will be excluded. ② **Type of design**: Randomized controlled trials from any country will be considered, provided they have been reported in English. ③ **Type of interventions**: Metformin alone or metformin supplemented with insulin. ④ **Type of outcomes**: The relevant outcomes will include birth weight, which is defined as a baby’s body weight at birth.

### Outcomes

The primary outcome is birthweight.

Secondary outcomes are Maternal Weight gain, Development of Gestational Diabetes, Development of hypertension/Preeclampsia Caesarian Section, Postpartum haemorrhage, Neonatal Hypoglycemia, Prematurity, Respiratory Distress, Macrosomia/Large for Gestational Age, Birth Trauma, Admission to NICU.

### Data collection and analysis

#### Study selection

Results of the search strategy will be imported into NoteExpress, and duplicate studies will be screened. According to the title and abstract, studies that do not meet the inclusion criteria will be excluded. Studies with uncertain eligibility will be screened after reading the full text. Suppose there is still uncertainty or disagreement after reading the full text. We will get in touch with the author to obtain relevant information before deciding or making further evaluations in the future selection process.Studies will be independently screened against the inclusion criteria by the reviewers. Any disagreement between the authors will be resolved through a consensus opinion among the reviewers. For practical reasons, none of the review authors will be blind to the journal titles, study authors, or institutions. We used a PRISMA flow diagram to record the results of the screening and selection processes. The original flow diagram is shown in figure 1.

**figure 1.**
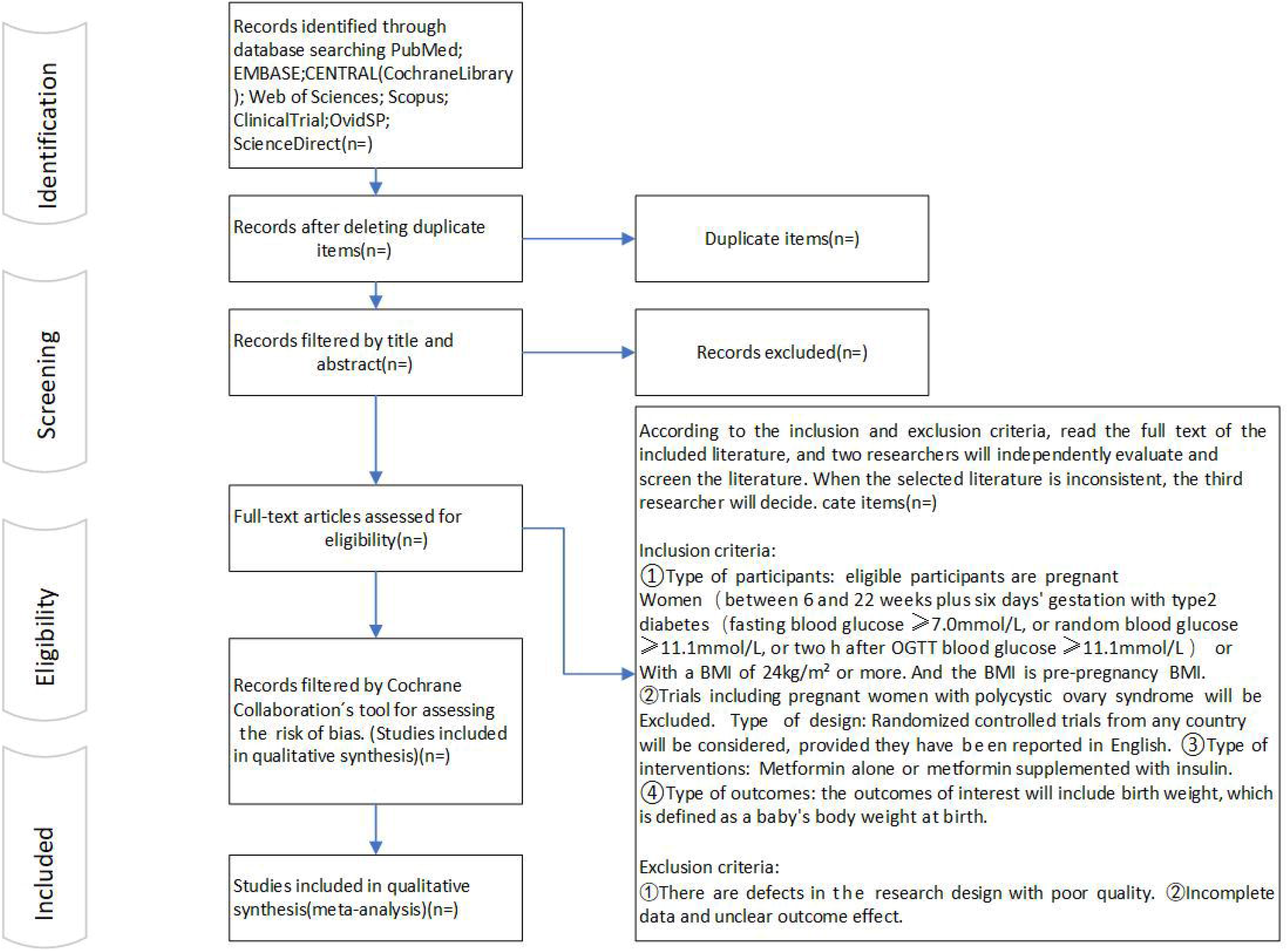

#### Data extraction

A data extraction form will be designed in advance, and the data will be extracted through Microsoft Excel 2019. The extracted data mainly include: (1) The basic information of the literature, including the title of the article, author, publication time, source of the literature, information of the evaluator, and others; (2) Main information of the study, including characteristics of the study object, study site, study design, data source, sample selection, data analysis, the background of research measures, research content, research methods, and measures to prevent bias; (3) Measurement of results, including follow-up time, loss of follow-up and withdrawal. For categorical variables, the total number of participants and event incidence in each group will be collected; for continuous variables, the number of participants, mean, standard deviation, or standard error will be collected. The data extraction form will be pre-tested before proper data extraction and refined based on the pre-test results.

In the data extraction process, if it is found that some reports belong to a single study, we will eliminate these repeated reports. If some critical data is not given in the study, the study’s authors will be contacted by email. If the author does not reply, we will contact the author every two days until the third time. Two independent investigators will extract the data, and a third independent investigator will cross-check all the data collected for any errors during data extraction. Any disagreement will be settled by consensus among the authors.

Other data items may be identified as necessary during the review process, any addition of data items will be clearly described, and the reason will be given.

#### Risk of bias in the individual study

We will use the Cochrane Collaboration tool for assessing the risk of bias, which covers: random allocation methods, allocation scheme concealment, use of the blind process, the integrity of result data, selective reporting of results, and additional sources of bias. We will assess each study on the above aspects and make a judgment of “low risk,” “high risk,” or “unclear.” “Unclear” refers to a lack of relevant information or uncertainty of bias. For such studies, we will contact the study authors for details. Two reviewers will make these judgments independently based on the criteria for judging the risk of bias. Any disagreement between the authors will be settled through a consensus opinion among the reviewers. Reviewers involved in this work will receive uniform training before commencement. The assessment results will be presented in tables and images and produced using RevMan(5.4.1; The Cochrane Collaboration).

### Assessment of heterogeneity

Clinical heterogeneity will be assessed by study participants, interventions, and outcome measures. Methodological heterogeneity will also be considered, including trial design and study quality. We used the Q test and the I^2^ test to assess heterogeneity. If the Q test result is P>0.10, heterogeneity is considered not statistically significant; the study is considered homogeneous, and the fixed-effect model will be adopted. Statistical heterogeneity is considered if the Q test result is P≤0.10. In this case, if I^2^ >50%, heterogeneity is considered to be too significant. After checking to confirm that the original data of each study and the method of data extraction are correct, meta-analysis will be abandoned, and only qualitative description will be carried out. If the heterogeneity test results P≤0.10, I^2^ ≤50%, the random effects model will be selected to estimate the combined effect size.

### Data synthesis

#### Data synthesis and meta-analysis approaches

The statistical analysis will be conducted using RevMan (5.4.1; The Cochrane Collaboration) and STATA17.

If the fixed-effects model is used, the dichotomous variables will be analysed using the Mantel-Haenszel method, and the continuous variables will be analysed using the inverse variance method. The DerSimonian-Laird method is used for all analyses if the random-effects model is used. We will conduct meta-analyses separately for the different effect measures (e.g., ORs, HRs, and risk ratios). The Z test will determine whether the combined effect size is statistically significant. If P≤0.05, the combined effect size is considered statistically significant. The forest plots will show all the content in the study’s statistical analysis.

#### Subgroup analysis

Subgroup analysis based on the following subgroups will also be considered appropriate:

▸ BMI before pregnancy (e.g. 24-30, 31-35, >35).
▸ Having type 2 diabetes or simply being overweight or obese. If other factors that warrant grouping are found in practice, subgroup analysis will also be performed after bunch. For continuous variables, the problem of grouping nodes will be carefully considered.

### Sensitivity analyses

When the heterogeneity between studies is too significant, sensitivity analysis will be performed to understand the source of heterogeneity. We will carefully interpret the sensitivity analysis results and further clarify the trustworthy source of the dispute.

### Publication bias

The possibility of publication bias will be investigated by visual assessment of a funnel plot and using Egger’s test, Begg’s test, and Harbord’s test if ten or more trials are pooled.

### Grading of evidence

The Grading of Recommendations Assessment, Developmentand Evaluation(GRADE) working group methodology will be used to classify the evidence quality into four grades: high quality, medium quality, low quality and very low quality, based on the design scheme of the included studies, the magnitude of the risk of bias, the consistency, indirection, accuracy of the findings and the possibility of reporting bias. The evaluation process and results will be presented in tabular form. The evaluations will be conducted independently by two reviewers. Any disagreement will be resolved by consensus among the authors.

### Patient and public involvement statement

This review was based on a collection of published literature and did not involve patient or public participation.

## Data Availability

All data produced in the present study are available upon reasonable request to the authors

## ETHICS AND DISSEMINATION

Only published literature will be used in our review. There will be no personally identifiable information and, therefore, no ethical issues. The results will be published in an international open-access journal.

## Author contributions

Conceptualisation: Kezhang Wang

Data curation: Kezhang Wang, Rui Pan

Formal analysis: Rui Pan, Yu Wang

Investigation: Yu Wang Methodology: Kezhang Wang, Rui Pan

Project administration: Kezhang Wang

Supervision: Kezhang Wang

Writing – original draft: Kezhang Wang, Rui Pan, Yu Wang

Writing – review & editing: Kezhang Wang, Rui Pan, Yu Wang

## Patient and public involvement

Patients and the public were not involved in the design, conduct, reporting, or dissemination plans of this research.

## Patient consent for publication

Not required.

## Provenance and peer review

Not commissioned; externally peer-reviewed.

